# Deconvolution of bulk endometrial tissue identifies cell type proportions and expression signatures associated with endometrial function and disease

**DOI:** 10.1101/2025.10.10.25337703

**Authors:** Monalisa Hota, Brett D. McKinnon, Peter A. W. Rogers, Grant W. Montgomery, Allan F. McRae, Sally Mortlock

## Abstract

**Background:** Human endometrium is a complex tissue lining the inside of the uterus and is essential for female fertility. It comprises several cell types, including endometrial stromal and epithelial cells, whose structure and function change across the menstrual cycle in response to steroid hormones. Although numerous bulk tissue studies have been carried out to investigate factors regulating gene expression in eutopic endometrium, these are complicated by variations in cell composition, cell specific expression profiles and the presence of endometrial pathologies. This study aims to investigate changes in the cellular and molecular environment of the endometrium across the menstrual cycle in women with and without endometriosis.

**Methods:** Using RNA sequencing data generated from the eutopic endometrium of 206 European women with and without endometriosis at different menstrual cycle phases, we estimated cell type proportions and cell type specific gene expression profiles using computational deconvolution methods.

**Results:** Cell type proportions varied across menstrual cycle phases and disease states. Women with endometriosis had lower proportions of luminal and ciliated epithelia at mid-secretory phase. We further analysed the estimated cell type specific gene expression profiles and identified differentially expressed (DE) genes within cycle phases between women with and without endometriosis in different cell types at the mid-secretory phase. Known genes associated with endometrial receptivity were DE including downregulation of *PTGS1* and upregulation of *POSTN* in stromal fibroblasts and glandular epithelia in women with endometriosis. DE genes were enriched in RNA metabolism and biogenesis pathways that can mediate fundamental mechanisms of cell proliferation and migration.

**Conclusions:** Our findings suggests that cell type deconvolution of RNA-sequencing data can improve our understanding of the role of cell type specific gene expression in endometrium, offering insights into potential disease relevant cell types and target genes.

## Background

The human endometrium is a complex tissue lining the inside of the uterus and is essential for female fertility. This tissue comprises several cell types, including endometrial stromal, epithelial and immune cells, whose structure, function and abundance change across the menstrual cycle in response to circulating steroid hormones (1, 2). Endometriosis is a benign oestrogen-dependent chronic gynaecological condition characterized by the growth of endometrial-like tissue outside the uterus in other body parts, commonly in the pelvic cavity(3). Endometriotic lesions can induce inflammation, scars and adhesions, and women with the disease can experience chronic pelvic pain and infertility(4). Whilst the mechanisms of endometriosis associated infertility are unknown, hypotheses include abnormal changes in the eutopic endometrium (inner lining of the uterus) that can also impact embryo implantation(5). Current evidence supports the role of retrograde menstruation in the aetiology of endometriosis in which endometrial stem and progenitor cells present in the eutopic endometrium flow back through fallopian tubes into the pelvic cavity(5). Therefore, studying the aberrations in eutopic endometrium is essential to understand the pathogenesis of endometrial disorders such as endometriosis and associated infertility.

Previous studies have investigated factors regulating gene expression in eutopic endometrium in women with and without endometriosis, these studies use biopsies containing multiple cell types, or “bulk tissue”, and thus may miss changes within individual cell types (6–9). As such, there is limited knowledge of the differences in cellular composition and cell type specific expression profiles in endometrium between individuals with and without disease throughout the menstrual cycle. Bulk tissue expression quantification methods like microarray and RNA-seq estimate the average expression of genes in a tissue, which is confounded by two factors of cellular heterogeneity: the relative abundance of each cell type and gene expression levels in these cell types(10). As a result, expression differences between samples with different conditions may arise due to variations in cell type proportions between conditions, actual gene expression changes, or a mixture of both(11). In recent years, single cell studies have greatly improved our understanding of cellular and molecular heterogeneity of endometrium and diseases(12–15). However, the high costs associated with these approaches have resulted in small sample sizes and are further constrained technologically, as they can only detect a limited number of genes. Alternatively, deconvoluting bulk tissue expression data can help reduce confounding effects caused by cell composition, while enabling the inclusion of larger sample sizes, thereby improving the power to study cellular and molecular heterogeneity within a tissue.

This study aims to investigate the dynamic changes in the cellular and molecular environment of the endometrium across different phases of the menstrual cycle in women with and without endometriosis. Cell type deconvolution methods were used to estimate cell type proportions and cell type specific gene expression from endometrium from 206 European women, one of the largest cohorts with gene expression data available for this tissue, including individuals with and without endometriosis at varying phases of the menstrual cycle(7). We show that the ability to deconvolute bulk endometrial expression data can improve our understanding of the role of cell type specific gene expression in endometrial function and disease.

## Methods

### 1. Data Collection

#### Endometrium bulk tissue RNA-seq data

Endometrial RNA sequencing data for this study was obtained from Teh et al.(16) and restricted to samples analysed in Mortlock et al.(7), which recruited 206 European women of reproductive age (143 endometriosis cases and 63 controls). The samples were collected from clinics at Royal Women’s Hospital (RWH) (184 gynaecology patients) and Melbourne IVF Clinic (IVF) (22 IVF patients) in Melbourne, Australia. Endometriosis status was confirmed surgically for the patients from RWH and self-reported for the patients from the IVF Clinic. Endometrial biopsies were collected from each woman and menstrual cycle phase was histologically confirmed by an experienced pathologist. The phases of the cycle were identified as menstrual (M), early-proliferative (EP), mid-proliferative (MP), late-proliferative (LP), early-secretory (ES), mid-secretory (MS) and late-secretory (LS). **Table 1** shows the number of endometriosis case and control samples collected at each phase.

**Table 1:** Bulk RNA-seq data sample information.

|  | RWH |  | IVF |  | Total |
| --- | --- | --- | --- | --- | --- |
| Endometriosis assessment | Surgically confirmed |  | Self-reported |  |  |
| Endometriosis status | Case<br>(n=135) | Control<br>(n=49) | Case<br>(n=8) | Control<br>(n=14) | 206 |
| Phase of menstrual cycle |  |  |  |  |  |
| Menstrual (M) (1) | 9 | 5 | 0 | 0 | 14 |
| Early-proliferative (EP) (2) | 3 | 2 | 0 | 0 | 5 |
| Mid-proliferative (MP) (3) | 56 | 15 | 0 | 1 | 72 |
| Late-proliferative (LP) (4) | 12 | 9 | 0 | 1 | 22 |
| Early-secretory (ES) (5) | 16 | 5 | 5 | 5 | 31 |
| Mid-secretory (MS) (6) | 24 | 7 | 3 | 7 | 41 |
| Late-secretory (LS) (7) | 15 | 6 | 0 | 0 | 21 |

#### Endometrium Single-cell RNA-Seq data

Several benchmarking studies have evaluated the performance of many popular partial, and complete deconvolution methods and suggested that partial deconvolution methods using single-cell data as a reference perform better than other methods(10, 17). As such, we used a single-cell RNA sequencing reference dataset available from Wang et al. containing endometrial superficial biopsies from 10 women without endometriosis, or uterine or pelvic pathology, and sequenced using the 10x Chromium system (GSE111976)(13). Annotation of donor samples and cell types was obtained from the supplementary table (GSE111976_summary_10x_day_donor_ctype.csv.gz). Samples were collected from these women on different days of the menstrual cycle (16, 17, 19, 20, 22, 23, 26). The phase of the menstrual cycle was assigned canonically using transcriptomic signatures by Wang et al. Two samples were obtained from the proliferative phase and the remining eight samples were identified across early, mid and late secretory phases.

### 2. Data analysis

#### Preparation of bulk RNA-seq data for deconvolution

Genes with expression values less than 10 counts (Counts Per Million, CPM <0.22) and expressed in less than 90% of the samples were considered as lowly expressed genes and removed. We corrected for batch (flow cell) using the ComBat_seq function from the sva v3.46.0 R package(18). Following TMM (Trimmed Mean of M-values) normalisation, counts per million (CPM) (edgeR v3.38.4 R package (19)) expression data was used for deconvolution analysis.

#### Preparation of single-cell RNA-seq reference dataset

Single-cell data were analysed and processed using the Seurat R package (4.3.0)(20). Cells with a detected number of genes less than 500 and high mitochondrial gene content (>15%) were removed, and then genes expressed in at least 1% of the cells were selected. Batch effect (donors) was corrected using the ComBat_seq function from the sva v3.46.0 R package(18). The data were normalized using the NormalizeData function in the Seurat package (normalization.method = "RC", scale.factor = 1e6). We selected 3000 variable features for further analysis. Clustering was done using the FindNeighbors (using 30 dimensions as a parameter) and FindClusters functions (using a resolution of 0.05) from the Seurat package. The cell types assigned by Wang et al. were matched to the identified clusters. Cells were removed if they were assigned as one cell type in the paper but clustered together with another cell type in our analysis. This reduced the number of cells to 54,223. From these 48,429 cells were from samples collected at the secretory phase and 5,795 cells were from proliferative phase samples. These cells covering the eight major cell types (stromal fibroblasts, glandular, luminal and ciliated epithelia, endothelia, smooth muscle cells, lymphocytes and macrophages) were retained for the deconvolution analysis. Marker genes that differentiate between cells in a cluster and all other cells (expressed in at least 25% of the cells in either group) were calculated using FindAllMarkers function with “wilcox” test. We further selected the top 300 genes (ranked by log fold change and FDR<0.05) from each cell type, totalling 2088 genes to estimate the cell type proportion using deconvolution analysis.

#### Estimation of cell type proportions

The CIBERSORTx(21) Fractions module was used to generate cell type proportions for 8 major cell types in the endometrium. The aforementioned single cell dataset consisting of 48,429 cells in secretory phase and 5,795 cells in the proliferative phase was used as the reference data set to deconvolute the corresponding menstrual phase bulk endometrium tissue samples. Batch correction (S mode) and 1000 permutations with other default options were used to run the deconvolution.

#### Differential cell type proportion analysis

Changes in cell type proportions among menstrual cycle phases in only control samples and between endometriosis cases and controls at each phase of the menstrual cycle were assessed using proportions obtained from CIBERSORTx. We first removed outlier samples within cases and controls in all phases and all cell types. Outlier samples were determined using Inter Quartile Range (IQR) method (samples that lie outside of the 25^th^ and 75^th^ quartiles +/- 1.5 * IQR were removed). P-values were calculated by performing two-sided Mann-Whitney U tests using wilcox.test() function in R. P-value < 0.05 was used as a cutoff to select cell types and phases where a significant change in proportion was observed. Furthermore, as a sensitivity analysis to adjust for the effect of age on the difference in proportions between endometriosis cases and controls, age was included as a covariate in the linear model.

#### Bulk differential expression (DE) analysis

We removed lowly expressed genes (genes expressed less than ∼10 counts in at least 90% of the samples) and raw gene counts were normalized for library size using the Trimmed Mean of M values (TMM) method in edgeR v3.38.4 R package(19). We used the limma v3.52.4 R package(22) to perform DE analysis. This analysis was done for 1) all endometriosis cases vs control samples, and 2) between cases and controls in each phase of the menstrual cycle. We fitted batch effects (flow cell and lane) as covariates for all analyses and in addition, fitted the phase of the menstrual cycle in model 1. The normalized counts were transformed using the voom() function in limma before fitting it to the linear model. Then eBayes method was used to contrast between the groups. The resulting P-values were adjusted using the Benjamini-Hochberg method. Significant differentially expressed genes were selected using FDR threshold < 0.05. To correct for the effect of cell type proportions we repeated the analysis described above and added the proportions as covariates in the linear model in addition to the other covariates.

#### Estimation of cell type specific expression

The bMIND(23) tool was used to estimate cell type specific gene expression profiles with Wang et al. single cell data as the reference dataset and cell proportions estimated using CIBERSORTx. Menstrual cycle phase (proliferative/secretory) specific single cell data was used to deconvolute the corresponding phase bulk samples. Both bulk and single cell data were log2-transformed prior to use.

#### Comparison of deconvoluted expression with single cell/single nuclei gene expression

We obtained publicly available single cell (sc) and single nuclei (sn) gene expression datasets from Human Endometrial Cell Atlas (HECA). HECA is a consensus cell atlas of the human endometrium containing transcriptomics and donor metadata information collected from published and newly generated datasets for around 300,000 single cells and 300,000 single nuclei (https://www.reproductivecellatlas.org/). In our study, we compared our deconvolution estimated expression data to Wang et al.(13) (used as a reference for deconvolution), Garcia-Alonso et al.(14) and Mareckova et al.(15) (sc and sn). We only used samples from these datasets from individuals without a history of endometriosis or using hormonal treatments. Phase-specific samples were used to compare the corresponding phase deconvoluted gene expression. In the sc/sn data sets, the mean expression for each gene was calculated from all cells (within a cell type, donors combined) within a phase (secretory/proliferative). For the cell type specific deconvoluted data, the mean expression for each gene was calculated from all samples within a menstrual phase. We then performed Spearman’s correlation to compare the datasets for 8 major cell types in the endometrium.

#### Cell type-specific DE and pathway analysis

To accommodate prior transformation of expression data (CPM normalized and log2-transformed prior to running bMIND), the differential expression analysis of estimated cell type-specific gene expression data was performed using the Wilcoxon rank sum test. We performed DE analysis in all cell types 1) between endometriosis cases vs control samples, and 2) between cases and controls in each phase of the menstrual cycle. The resulting P-values were adjusted using the False Discovery Rate (FDR) method. Significant differentially expressed genes were selected with FDR threshold < 0.05. To further assess the impact of age on the gene expression differences between endometriosis cases and controls within menstrual phases, a sensitivity analysis was conducted by incorporating age as a covariate in the linear model.

Pathway analysis was performed in Metascape (https://metascape.org) v3.5.20240101(24) using cell type specific upregulated (UP) or downregulated (DOWN) genes (FDR threshold < 0.05) in endometriosis cases. Number of DE genes to perform the pathway enrichment analysis in each category are as follows: Stromal fibroblasts - UP n=325, DOWN n=783, glandular epithelia - UP n=198, DOWN n=534, Luminal epithelia - UP n=1281, endothelia - DOWN n=327.

## Results

### Study Overview

Our analysis approach, summarized in Figure 1, integrated single-cell and bulk RNA-seq data to investigate cell type specific changes in the endometrium across the menstrual cycle and between women with and without endometriosis. Using CIBERSORTx and bMIND, we deconvoluted bulk RNA-seq data from 206 individuals into cell type proportions and gene expression profiles for eight major endometrial cell types (stromal fibroblasts, glandular, luminal and ciliated epithelia, endothelia, smooth muscle cells, lymphocytes and macrophages). Phase specific single-cell data were used as references to account for significant transcriptional differences between proliferative and secretory phases. We compared cell type proportions across menstrual cycle phases and between endometriosis cases and controls, incorporating these proportions as covariates in differential gene expression analyses. Finally, we identified phase specific and cell type specific differentially expressed genes and enriched pathways.

**Figure 1.**
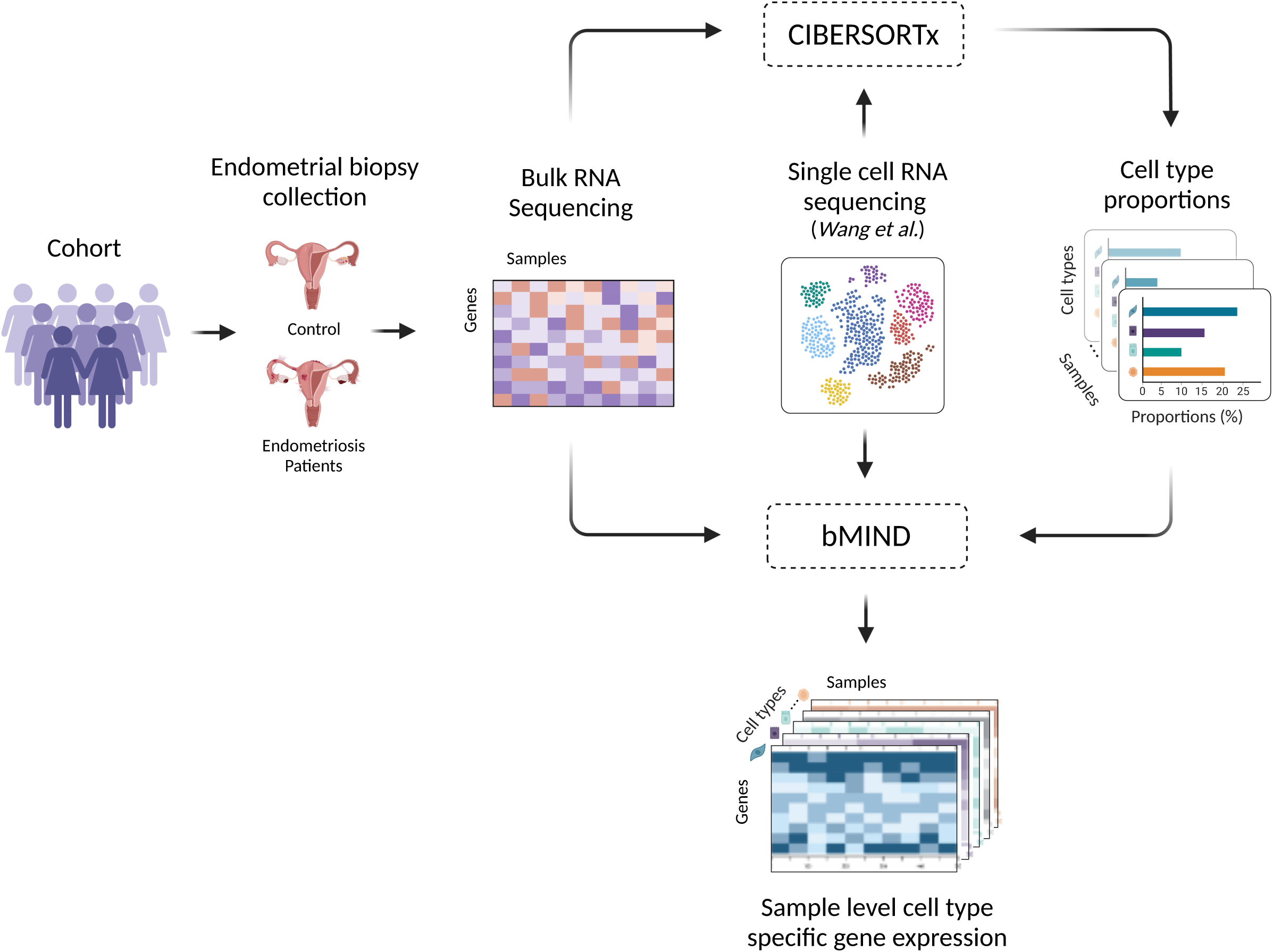
Analysis overview. Bulk RNA samples from endometrial biopsies were deconvoluted into individual cell types allowing comparison of cell type proportions and cell specific gene expression across the menstrual cycle and with disease.

### Changes in cellular composition across the menstrual cycle

The cellular composition of endometrium varies across the menstrual cycle(15, 25). Using CIBERSORTx we estimated cell type proportions in each sample. The mean estimated proportions of the 8 major cell types in the endometrium across the phases of menstrual cycle are shown in Figure 2a (Additional file 2: Table S1). Stromal fibroblasts are the most abundant, with the mean proportion ranging from 67% in the menstrual phase to 45% in the early secretory phase. Overall, stomal fibroblast proportions were higher in proliferative phases compared to secretory phases. Meanwhile, the proportions of the glandular epithelium were lower in the early proliferative phase and gradually increased through the menstrual cycle, ranging from 5% in the menstrual phase to 35% in the mid-secretory phase. The remaining cell types made up proportions ranging from 0.3% to 9% (0.5% to 9% for luminal epithelium; 1% to 4% for ciliated epithelium; 2% to 8% for endothelium; 0.3% to 5% for smooth muscle, 1% to 8% for lymphocytes and 0.7% to 3% macrophages) (Figure 2a, Additional file 2: Table S1).

**Figure 2.**
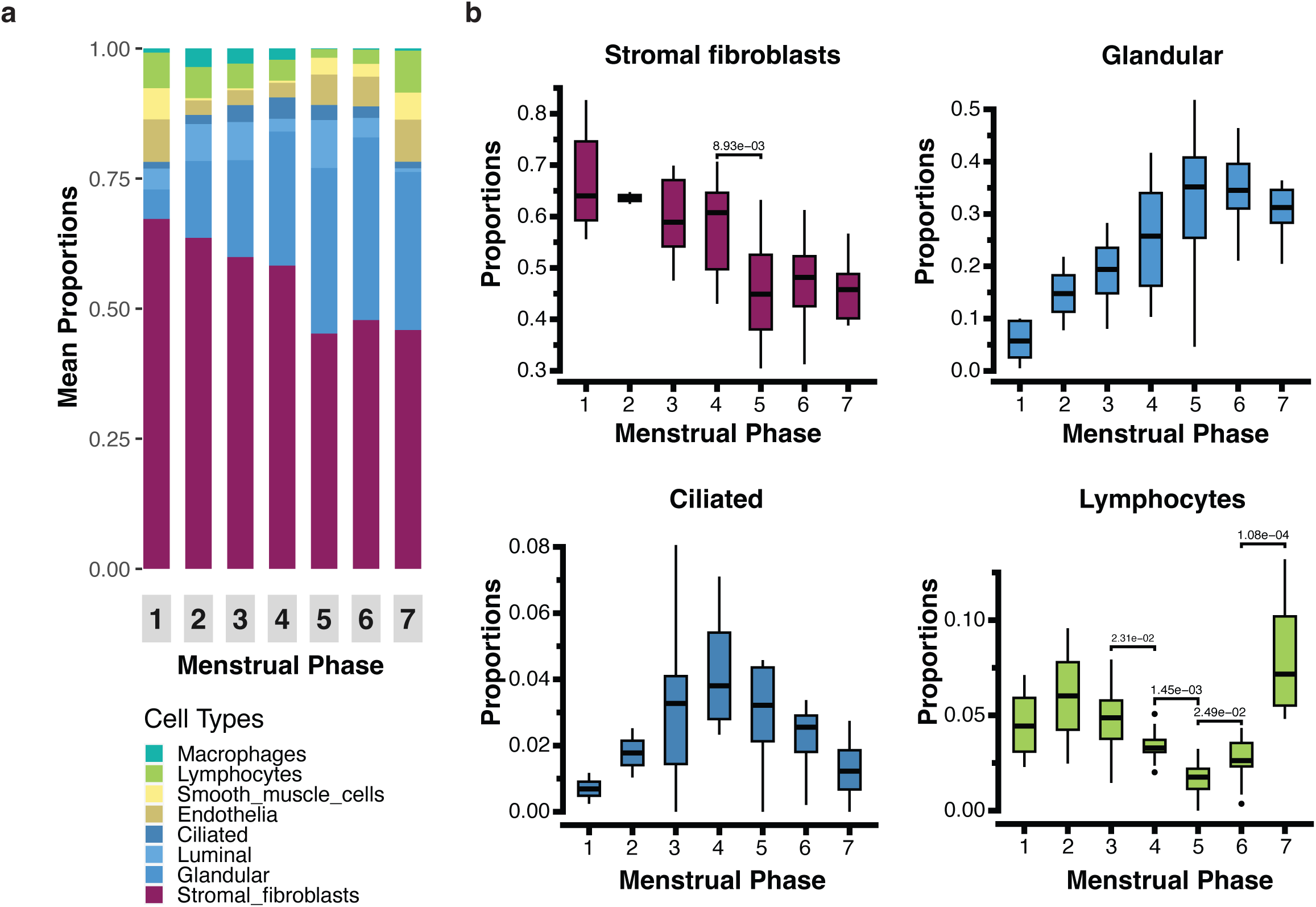
Cell type proportions of control samples across menstrual phases. (**a**), Mean proportion of each estimated cell type in control samples in the seven menstrual cycle phases. The cycle phases numbered 1-7 are defined as following: 1- Menstrual, 2- Early-proliferative, 3- Mid-proliferative, 4- Late-proliferative, 5- Early-secretory, 6- Mid-secretory and 7- Late- secretory. (**b**), Boxplots showing the distribution of estimated proportions for each cell type at each menstrual cycle phase. The phases of the menstrual cycle numbered 1-7 as defined in a. Significant changes in the cell type proportion between consecutive menstrual cycle phases are annotated with p-values highlighted in the plot.

All cell types showed significant changes (Kruskal-Wallis rank sum test p-value < 0.05) in estimated proportions throughout the menstrual cycle (Additional file 2: Table S2). Comparisons of cell type proportions between different phases of the menstrual cycle revealed significant changes (p-value<0.05) in the proportion of some cell types between one phase of the menstrual cycle and the next phase (Figure 2b, Additional file 1: Figure S1). Most significant changes in proportions were observed across different phases of secretory endometrium as shown in Figure 2b and Additional file 1: Figure S1. Stromal fibroblast proportions significantly decreased from late-proliferative (LP) to the early-secretory (ES) phase. Statistical tests showed a significant change in glandular and ciliated epithelia proportions across the menstrual cycle. However, we did not identify any significant difference in proportions between any two consecutive phases, which could be due to sample level variation. Luminal epithelium proportions significantly increased from LP to ES and decreased throughout the secretory phases (Additional file 1: Figure S1). Endothelia cell type proportions significantly increased from LP to ES then increased again in mid secretory (MS) to late secretory (LS) phase (Additional file 1: Figure S1). Similar changes in proportion are observed with smooth muscle cells. Proportions of lymphocytes decreased from mid proliferative (MP) to LP to ES, then increased through the secretory phases (Figure 2b). Macrophage proportions significantly decreased from LP to ES then increased across the secretory phases (Additional file 1: Figure S1). Overall, we observe dynamic changes in the proportions of cell types in endometrium across the menstrual cycle.

### Difference in cell type proportions between endometriosis cases and controls

In addition to assessing changes in cell type proportion in controls across phases of the menstrual cycle, we also tested for differences in cell type proportions between samples from endometriosis cases and controls within each phase. We observed significant (p-value < 0.05) differences in cell type proportions between cases and control for luminal and ciliated epithelia, endothelia and smooth muscle cells in the mid secretory phase, for stromal fibroblasts, endothelia, glandular and ciliated epithelia in the menstrual phase, and only for smooth muscle cells in the late proliferative phase (Figure 3) (Additional file 1: Figure S2). A study by Loid et al., demonstrated that older women have a higher proportion of ciliated epithelial cells in their endometrium in secretory phase. Accounting for age in the model, we show differences between endometriosis cases and controls remained consistent (Additional file 2: Table S3) (26). Differences observed in the menstrual phase of the cycle should be interpreted with caution given the large variation observed in this phase and the small sample size. A significant difference was observed in luminal epithelia at the mid secretory phase where the cell type proportion is lower in case samples compared to control samples (Figure 3). Ciliated cell proportions were also lower in cases compared to controls at the mid-secretory phase. In contrast, the proportions of endothelia and smooth muscle cells were higher in cases compared to controls. Since cell proportions must collectively sum to 100%, an observed increase in one cell type implies a corresponding decrease in at least one other cell type. As a result, changes should be interpreted within this compositional context. No significant differences in cell type proportions between endometriosis cases and controls were detected in any other phases in any cell type.

**Figure 3.**
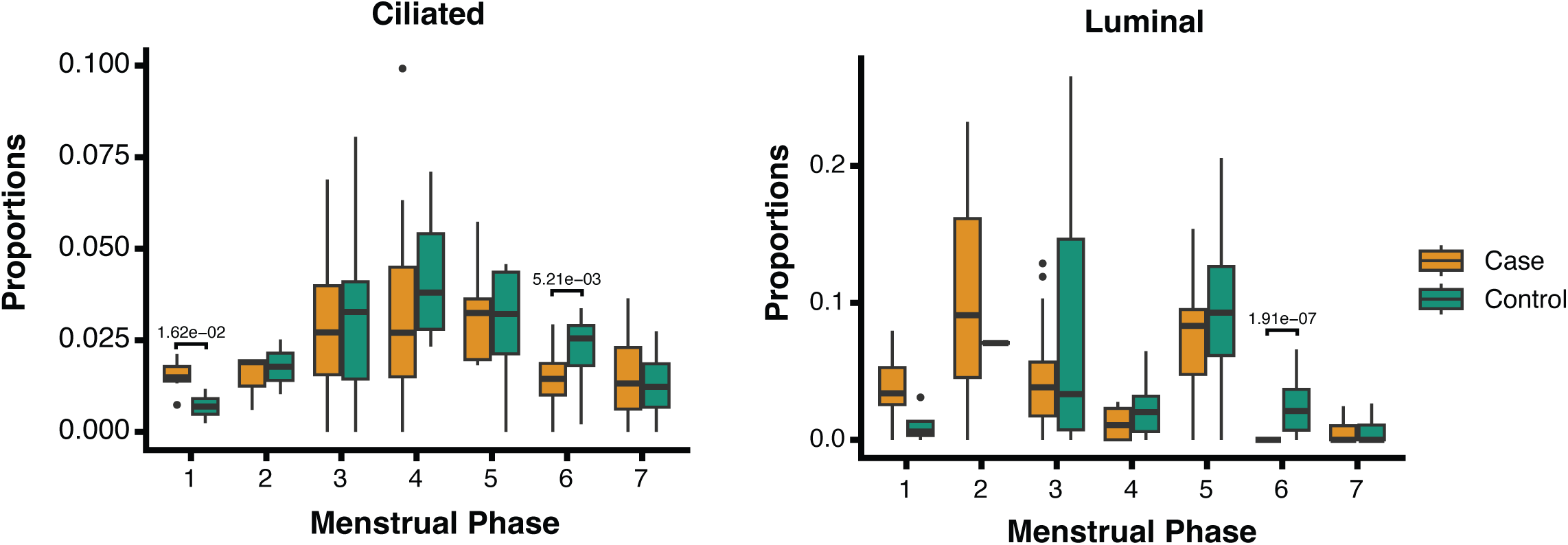
Difference between the proportions of ciliated and luminal cell types between cases (green) and controls (orange) in each of the seven menstrual cycle phases. The phases of the menstrual cycle numbered 1-7 are defined in Figure 2a.

### Differential expression following correction for cell type proportions

Differential expression (DE) analysis was performed between all endometriosis cases and controls and between cases and controls at each phase of the menstrual cycle using the limma package. We did not identify any DE genes when comparing all cases and controls however, we identified 42 differentially expressed genes (FDR < 0.05) between cases and controls when the analysis was restricted to the mid secretory phase (Additional file 2: Table S4).

Traditional bulk tissue DE analysis doesn’t consider cell type proportions, as a result DE genes observed in such analyses can be false positives. To account for differences in cell type proportions among samples we used our estimated cell type proportions as covariates in the limma model. After correcting for proportions, we did not observe any significant DE genes between all cases and all control samples, consistent with the previous analysis (7). Only PDE3A was upregulated with FDR threshold < 0.05 between cases and controls within the mid-secretory phase (phase 6) (Additional file 2: Table S4), compared to 42 in the DE analysis without proportion correction.

### Cell type specific expression estimates

Cell type proportions estimated by CIBERSORTx and single cell data from Wang et al. were included as reference alongside bulk expression data for 13,680 genes to obtain cell type-specific gene expression profiles using the bMIND deconvolution tool. To examine whether estimated cell type specific expression profiles could detect cell type specificity despite being derived from bulk tissue samples, we performed principal component analysis (PCA) of the estimated cell type and bulk gene expression profiles. PCA results showed that the major source of variation is contributed by the cell types (Figure 4a).

**Figure 4.**
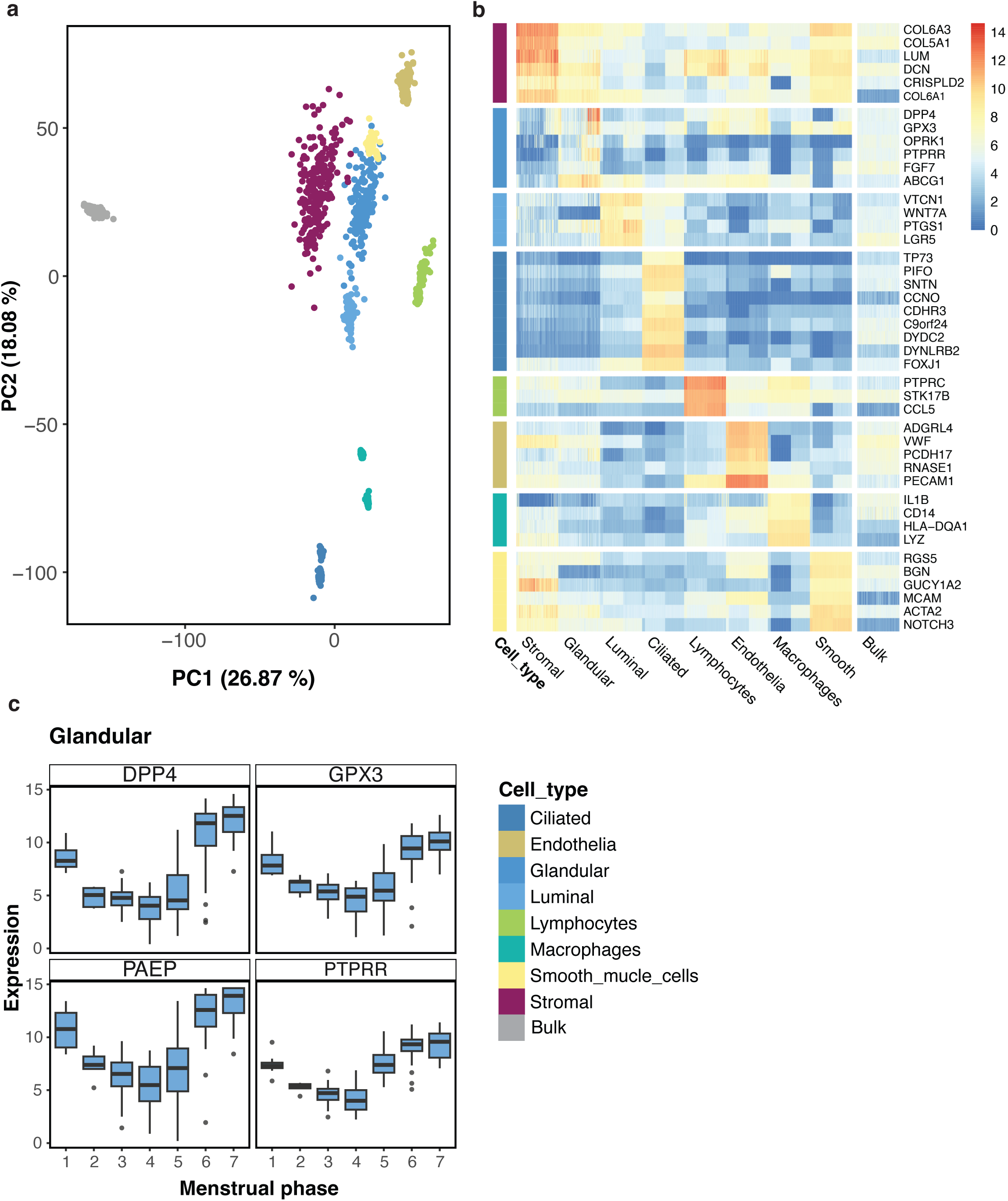
(**a**) PCA of bMIND estimated cell type expression and bulk tissue expression. The colour of the dots represents different cell types and bulk tissue samples. (**b**) Heatmap plot of cell type specific marker genes. As expected, the cell type marker genes are most highly expressed in their aligned tissue. (**c**) bMIND estimated gene expression levels of genes which are expressed in higher levels in the secretory phase compared to proliferative in the glandular epithelial cell type.

Next, we examined the expression levels of known marker genes for each cell type obtained from Wang et al. The heatmap plot (Figure 4b) shows that cell type specific marker genes are highly expressed in the corresponding cell type compared to the other cell types and are arbitrarily expressed in the bulk tissue. These results suggest that computationally derived cell type specific gene expression estimates capture cell type specific signals that are consistent with known expression levels of cell type specific marker genes.

In addition to checking cell type marker expression, we also checked the expression levels of menstrual cycle phase marker genes (Additional file 1: Figure S3). Figure 4c shows the expression levels of the genes that are highly expressed in secretory phase in glandular epithelia cell type compared to proliferative phase, which suggests our deconvoluted expression data has captured menstrual cycle phase specific differences in appropriate cell types.

### Comparison of deconvoluted and sc/sn gene expression

We compared deconvolution estimated gene expression data with publicly available sc/sn expression data from Human Endometrial Cell Atlas (HECA) (15). A strong correlation between deconvoluted estimates and sc/sn data in stromal fibroblasts is observed in both the proliferative (R2: 0.64-0.75) and secretory phase (R2: 0.58-0.71) as shown in Additional file 1: Figure S4. There was a moderate correlation in the proliferative phase (R2: 0.37 – 0.5) and a strong correlation in secretory phase (R2: 0.61 -0.75) observed for glandular epithelia. The full list of correlations in different cell types in the proliferative and secretory phase are shown in Additional file 2: Table S5. Overall, we observed moderate to strong correlation between deconvolution estimates and sc/sn datasets. The correlation between different single cell datasets ranged from 0.7 - 0.98 depending on the cell type, which is expected given we obtained the sc datasets from HECA atlas, where all these datasets are harmonized and integrated together to adjust for dataset level variation. The correlation between the single cell datasets and single nuclei dataset were much lower ranging from 0.27 in lymphocytes to 0.6 in stromal fibroblasts (Additional file 2: Table S5). We observed higher correlation among the higher abundant cell types compared to lower abundant cell types, likely due to more variability in estimates in the lower abundant cell types resulting from a smaller number of cells in the sc/sn data.

### Cell type specific DE analysis between cases and controls at different phases of the menstrual cycle

Lastly, differential expression analysis between all endometriosis cases (143 samples) and all controls (63 samples) for 8 major cell types was performed using the Wilcoxon rank sum test. No genes were significantly (FDR < 0.05) differentially expressed in any of the cell types. This could be a result of high variation in gene expression across the menstrual cycle(15, 27). As such, we performed DE analysis between cases and controls within each phase of the menstrual cycle for all cell types. Out of 7 phases of menstrual cycle, we only observed DE genes in the mid-secretory (phase-6) phase in stromal, epithelial and endothelial cell types. Based on a significance threshold of FDR < 0.05 we observed 1,301 DE genes in luminal epithelia, 1,108 DE genes in stromal fibroblasts, 732 DE genes in glandular epithelia, 340 DE genes in endothelia and only 7 DE genes in ciliated epithelia (Additional file 2: Table S6). The DE genes with FDR < 0.05 and absolute log2 Fold change (log2FC) >1 are shown in the volcano plots (Figure 5). DE genes with a log2FC>1 were only observed in glandular epithelia and stromal fibroblasts. We observed that 9 DE genes (*STAR*, *OMG*, *MT1G*, *MT1X*, *FABP5*, *PTGS1, LEFTY1*, *BRINP1*, *FERMT1*) were down-regulated in both cell types and 5 DE genes (*THBS1*, *OLFML2A*, *SGIP1*, *CELF2*, *ABCA9*) were up-regulated in both cell types. Results remained consistent when age was included as covariate, though statistical power decreased due to the additional degree of freedom. Nonetheless observed fold changes were highly correlated (Additional file 1: Figure S5).

**Figure 5.**
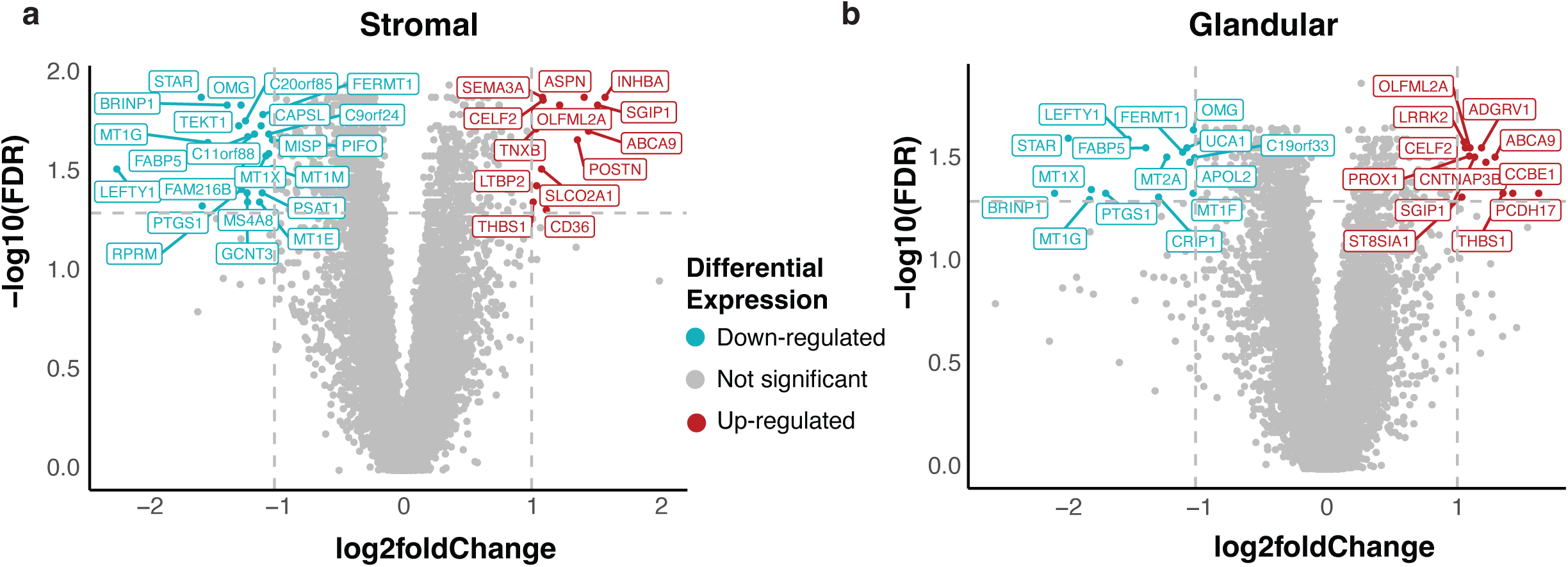
Fold change and FDR volcano plot for stromal fibroblasts and glandular epithelia in mid-secretory phase. The significant DE genes with FDR < 0.05 and absolute log2 Fold change >1 are labelled in the plot. The up-regulated genes are coloured red and down-regulated genes are coloured blue.

Endometrium becomes receptive to blastocyst implantation in the mid-secretory phase which is called Window Of Implantation (WOI)(2). Hence, we investigated if any of the DE genes observed are genes known to be associated with receptivity. Using a selection of previously published genes associated with receptivity (6, 28), we observed that *POSTN* was upregulated (log2FC >1) in endometriosis cases in stromal fibroblasts and glandular epithelia, and *STAR*, *PTGS1*, *MT1G*, *RPRM*, and *MT1X* were downregulated (log2Fc < -1) in endometriosis cases in stromal fibroblasts. *STAR*, *MT1G*, *MT1X*, *PTGS1*, *MT2A*, *APOL2* and *MT1F* were also downregulated (log2Fc < -1) in endometriosis cases in glandular epithelia compared to controls. Receptivity-associated genes in all cell types that are differentially expressed between endometriosis cases and controls with FDR < 0.05 are listed in Additional file 2: Table S6 with their corresponding log2 fold changes.

To obtain insights into biological pathways enriched with significant DE genes between endometriosis cases and controls in the stromal fibroblasts, glandular, luminal epithelia and endothelia at mid-secretory phase, we performed functional enrichment analysis using Metascape. The top 20 gene ontology terms enriched for upregulated and downregulated genes in endometriosis cases are shown in Additional file 1: Figure S6. The full pathway analysis results are available in Additional file 2: Table S7-12. Pathway analysis highlighted pathways enriched for downregulated genes in stromal fibroblasts and glandular epithelia, i.e. “metabolism of RNA”, “translation”, “regulation of cellular response to stress” etc. Enriched pathways on upregulated DE genes show enrichment in “positive regulation in cell migration”, “sensory system development”, “regulation of cell-substrate adhesion”, cell-cell adhesion”, “gland development” etc. pathways in stromal fibroblasts and glandular epithelia.

## Discussion

The complexity of the human endometrium, including its dynamic cellular composition and molecular heterogeneity across the menstrual cycle(1, 2), makes it increasingly difficult to understand and study endometrial disorders such as endometriosis. To address this, we applied advanced cell type deconvolution methods to bulk RNA-seq data from eutopic endometrial tissue collected at different menstrual cycle phases in women with and without endometriosis. By integrating gene expression data from a large bulk RNA-seq dataset with cell type-specific signatures derived from single-cell RNA-seq data, we investigated the cellular and molecular heterogeneity of the endometrium and its alterations in endometriosis. Our findings reveal significant differences in cell type proportions and gene expression profiles during the mid-secretory phase, including reduced proportions of luminal and ciliated epithelia, differential expression of endometrial receptivity-associated genes, and enrichment of pathways involved in RNA metabolism, cell proliferation, and migration.

Deconvolution of bulk endometrial RNA-seq data allowed us to identify changes in cellular proportions across the menstrual cycle in a large number of samples which were consistent with the proportions observed in recent single-cell and spatial transcriptomic studies(14, 15). Kruskal-Wallis rank sum tests showed that proportions of all cell types changed dynamically across the menstrual cycle (p-value < 0.05). The increase in the proportion of ciliated and glandular epithelium from early proliferative to secretory phases and sustained proportion of glandular epithelium across the secretory phase is consistent which what has been observed histologically(29, 30). In addition, comparison of cell type proportions in consecutive phases, showed significant (p-value <0.05) difference in proportions between the late proliferative phase to early secretory, and mid-secretory to late secretory phase, in endothelial, smooth muscle cells and immune cells. Cell type proportions boxplots (Figure 2b) showed changes in proportions of epithelial, stromal and immune cells throughout the menstrual cycle. However, for glandular and ciliated epithelia, we didn’t observe a statistically significant difference in proportion between any two consecutive phases. Since cellular changes across the cycle are likely to occur on a continuum, we expect sample level variation within each phase, given that biopsy samples were collected on different days of the menstrual cycle and individuals have varying cycle lengths(31). These results suggest that computationally derived cell type proportions are consistent with expected biology even in a dynamic tissue such as the endometrium.

In addition to cell type proportion changes across menstrual cycle, we also identified proportion changes between endometriosis cases and controls within cycle phases (Figure 3). We observed a decrease in the proportions of luminal and ciliated epithelia in endometriosis cases in the mid-secretory phase. In the mid-secretory phase, the endometrium becomes receptive to embryo implantation(2). Endometrial glands extend towards the uterine cavity and are lined by a layer of luminal and ciliated epithelial cells which play key roles in preparing the endometrium for embryo implantation (14, 15, 32, 33). Approximately 50% of the women with endometriosis experience impaired fertility(34, 35). Therefore, the observed changes in epithelial cell proprotions during the mid-secretory phase, and their potential influence on receptivity and endometriosis-associated infertility, warrents further investigation. Differential expression analysis of bulk RNA-seq data between cases and control in the mid-secretory phase identified 42 significant DE genes (7). However, after correcting for estimated cell type proportions for major cell types as covariates these genes did not pass the FDR threshold. As bulk tissue expression is the sum of the abundances of all cell types and gene expression levels for these individual cell types, the above observation suggests that DE genes observed before correction may be the consequence of the cell type proportion changes, not gene expression changes.

This study highlights the ability of computationally estimated cell type proportions and cell type specific expression estimates generated from deconvolution of bulk endometrium expression data can capture cell type and menstrual phase specific signals (Figure 4a b c). Comparison of the deconvoluted estimates to sc/sn datasets showed moderate to strong correlation. Sn/sn datasets are sparse and a lot of dataset level variation exists among them. Hence observed correlation from the datasets are in expected range based on knowledge of the technologies. Results from DE analysis of cell type specific expression estimates at mid-secretory phase identified upregulated and downregulated genes in endometriosis cases in stromal fibroblasts, glandular, luminal and ciliated epithelia and endothelia. Amongst the DE genes identified, 33 genes (Additional file 2: Table S6) were previously reported to be associated with endometrial receptivity(6) across different cell types, suggesting that changes in expression levels of these genes at the receptive phase of the menstrual cycle may contribute to endometriosis associated infertility. One such receptivity-associated gene, *PTGS1* (Prostaglandin-Endoperoxide Synthase 1) was downregulated in endometriosis cases (Log2FC: -1.69 in glandular epithelia, -1.56 in stromal fibroblasts and -0.3 in endothelia). *PTGS1* and 2 (COX-1 and -2) control synthesis of Prostaglandins (PGs) and are involved in developmental regulation during peri-implantation and dysregulation has been associated with recurrent implnantation failure (36–41). PGs involved in decidualization are generated by *PTGS1* and endometrial receptivity can be affected by change in the PG synthesis and metabolism(36). Hence, down-regulation of *PTGS1* in different cell types in the receptive phase in endometriosis patients could affect blastocyst implantation. Alternatively, *POSTN* (Periostin), another receptivity-associated gene, was upregulated in stromal fibroblast (Log2FC: 1.36) and glandular epithelia (Log2FC: 0.91) in endometriosis cases compared to the controls. This is a cell adhesion protein which is shown to enhance migration, invasion and adhesion of stromal cells through Integrin-Linked Kinase 1/Akt Signaling Pathway(42) and epithelial cells through Epithelial-Mesenchymal Transition(43). Significantly higher expression of *POSTN* has been previously observed in eutopic endometrium of endometriosis patients (44). In addition, Morelli et al. (45) suggested that *POSTN* is involved in embryo– endometrial cross talk at implantation and should be considered as biomarker of endometrial receptivity. Potential roles of the DE genes in endometriosis and endometriosis-associated infertility warrant further investigation.

Neuroangiogenesis is one of the theories proposed to explain development of endometriosis which hypothesises that the factors regulating angiogenesis and redistribution of nerve fibers play a critical role in the pathophysiology of endometriosis(46). These regulatory factors may also affect establishment of the lesions in the peritoneal cavity. *SEMA3A* (Semaphorin 3A) is a member of family of factors regulating nerve distribution, growth and regulation of angiogenesis. A recent study by Yang et al.(47) using knocked down *SEMA3A* suggested that hypoxia plays an important role in pathophysiology of endometriosis promoting proliferation and migration of endometrial stromal cells by upregulating *SEMA3A.* In our data, we observed significant upregulation of *SEMA3A* in stromal fibroblasts (Log2FC: 1.08), glandular epithelia (Log2FC: 0.43) and endothelia (Log2FC: 0.23) in endometriosis patients at mid-secretory phase. Further investigation is required to clarify the functional consequences of increased expression of this gene during the mid-secretory phase of the eutopic endometrium. While it may influence ectopic cells via retrograde menstruation and thereby affect the risk of developing endometriosis, this hypothesis remains speculative. In addition, in our pathway analysis of upregulated genes in stromal fibroblasts and glandular epithelia, we observed significant enrichment of pathways such as “sensory system development” and “cell-cell adhesion”, which suggests that upregulation of group of genes in addition to *SEMA3A* may be involved in promoting lesion formation in peritoneal cavity.

Genome-wide association studies (GWAS) have identified genetic risk loci associated with endometriosis(48) and infertility(49), but the functional relevance of genes within these loci remains unclear, prompting us to investigate if genes in these loci were differentially expressed in our deconvoluted dataset. Eight genes mapped to endometriosis risk loci were differentially expressed in endometriosis cases at mid-secretory phase including, *GREB1*, *CD109*, *KDR*, *DNM3*, *FAM120B*, *KCTD9*, *ASTN2* that were upregulated in cases and *HOXA10* that was downregulated (Additional file 2: Table S6). Similarly, *GREB1*, *PROX1*, *NCKAP5* and *ZEB2* that map to infertility risk loci are upregulated cases. *GREB1* (Growth Regulating Estrogen Receptor Binding 1) is known to participate in normal endometrial function and receptivity as well as facilitates the progression of estrogen-driven endometriosis (50). Dysregulation of this gene in endometriosis cases at mid-secretory phase of the menstrual cycle further supports its role in endometriosis associated infertility. These findings provide valuable insights into the regulation and potential roles of genes mapped to endometriosis and infertility risk loci, suggesting that their differential expression in endometriosis may contribute to disease pathogenesis and infertility, whether driven by genetic regulation or other molecular mechanisms. There are inherent limitations associated with studying a dynamic tissue like endometrium, such as individual level and cycle level variation in cell differentiation, gene expression and menstrual cycle length. Large gene expression changes across the mid-secretory phase suggest that differences in cycle day among samples within this stage may also contribute to the observed variability (16). Cell type deconvolution from bulk RNA-seq is limited by its dependence on reference datasets, which may be incomplete or biased, and marker gene selection, which can be unreliable. Resolution is constrained, making it difficult to distinguish closely related cell types, especially with variability in cell states. Similarly, rare cell types are challenging to estimate accurately, and dominant cell populations may overshadow their signals. These challenges collectively impact the accuracy and robustness of deconvolution methods. To assess the reliability of our estimates we have compared our gene expression levels to other publicly available endometrium single cell and single nuclei datasets and critically evaluated our findings with existing knowledge in endometrial biology. Further validation in large independent cohorts with detailed menstrual cycle staging and high-resolution single-cell studies is reccomended to confirm these observations.

## Conclusions

In conclusion, our study highlights the power of computational cell type deconvolution in uncovering the complex cellular and molecular landscape of the human endometrium. By analysing RNA sequencing data from women with and without endometriosis across different menstrual cycle phases, we demonstrated significant variations in cell type proportions and identified key differentially expressed genes associated with endometrial receptivity. Bulk RNA-seq deconvolution is a scalable and cost-effective approach that leverages existing datasets to estimate cell type composition, enabling the study of large sample cohorts and subtle cellular changes. These findings provide insights into endometrial biology and its link to endometriosis, emphasizing the importance of integrating computational approaches to refine our understanding of cell type specific gene expression in complex tissues.

## Data Availability

The bulk RNA-seq data (16) are publicly available at GEO under accession number GSE234354. The single cell RNA data (13) are publicly available at GEO under accession number GSE111976. Other datasets supporting the conclusions of this article are included within the article and its additional files.

https://www.ncbi.nlm.nih.gov/geo/query/acc.cgi?acc=GSE234354

https://www.ncbi.nlm.nih.gov/geo/query/acc.cgi?acc=GSE111976

## Declarations

### Ethics approval and consent to participate

The study was approved by the Royal Women’s Hospital Human Research Ethics Committee (Projects 11-24 and 16-43), the Melbourne IVF Human Research Ethics Committee (Project 05-11) and the University of Queensland. Informed consent was obtained from all participants.

### Consent for publication

Not applicable

### Competing interests

The authors declare that they have no competing interests.

### Funding

M.H is funded by Research Higher Degree Scholarship, The University of Queensland. AFM is funded by an Australian Research Council Future Fellowship (FT200100837). SM is funded by a National Endometriosis Clinical and Scientific Trials Fellowship (UNSW-RG223907).

### Authors’ contributions

S.M, M.H, A.F.M and G.W.M designed the study with input from the other authors. Data analysis was performed by M.H which was interpreted by all authors. M.H, S.M and A.F.M wrote the manuscript with input from all other authors. The final report has been critically revised and approved by all authors.

## Acknowledgements

We gratefully acknowledge all study participants at the Royal Women’s Hospital and Melbourne IVF Clinic, as well as the authors who contributed to Teh et al. (16) dataset used in this study.

## Additional Files

Additional file 1

File format: .pdf

Supplementary figures.

Additional file 2

File format: .xls

Supplementary tables.

## Notes

### Competing Interest Statement

The authors have declared no competing interest.

### Author Declarations

The study used ONLY openly available human data that were originally located at GEO.

