## Supplementary Figures for "Deconvolution of bulk endometrial tissue identifies cell type proportions and expression signatures associated with endometrial function and disease"

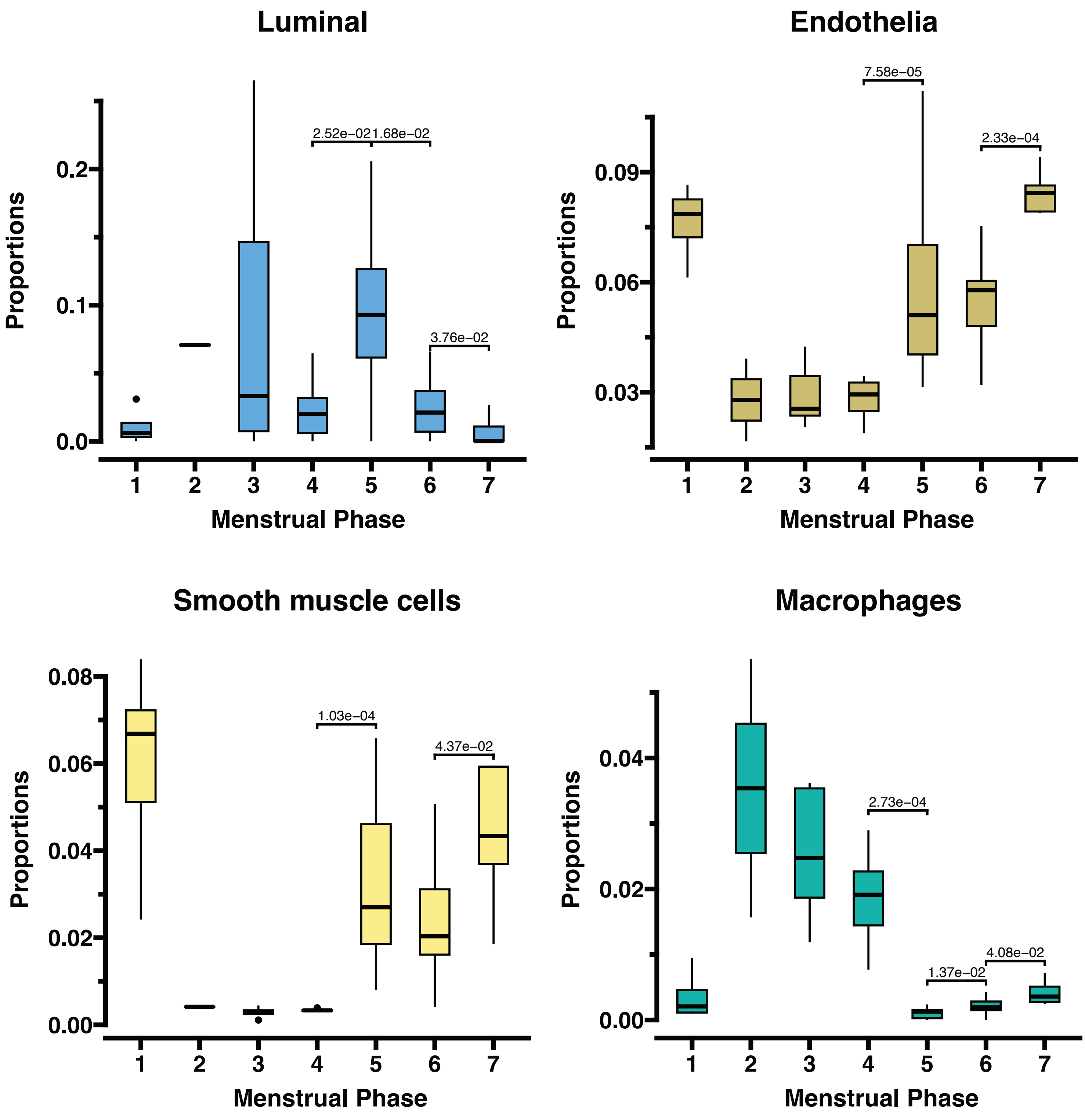


**Figure S1 -** Cell type proportions of control samples across menstrual phases. The menstrual cycle phases are defined in Figure 2a. Significant changes in the cell type proportion between consecutive menstrual cycle phases are annotated with p-values highlighted in the plot.

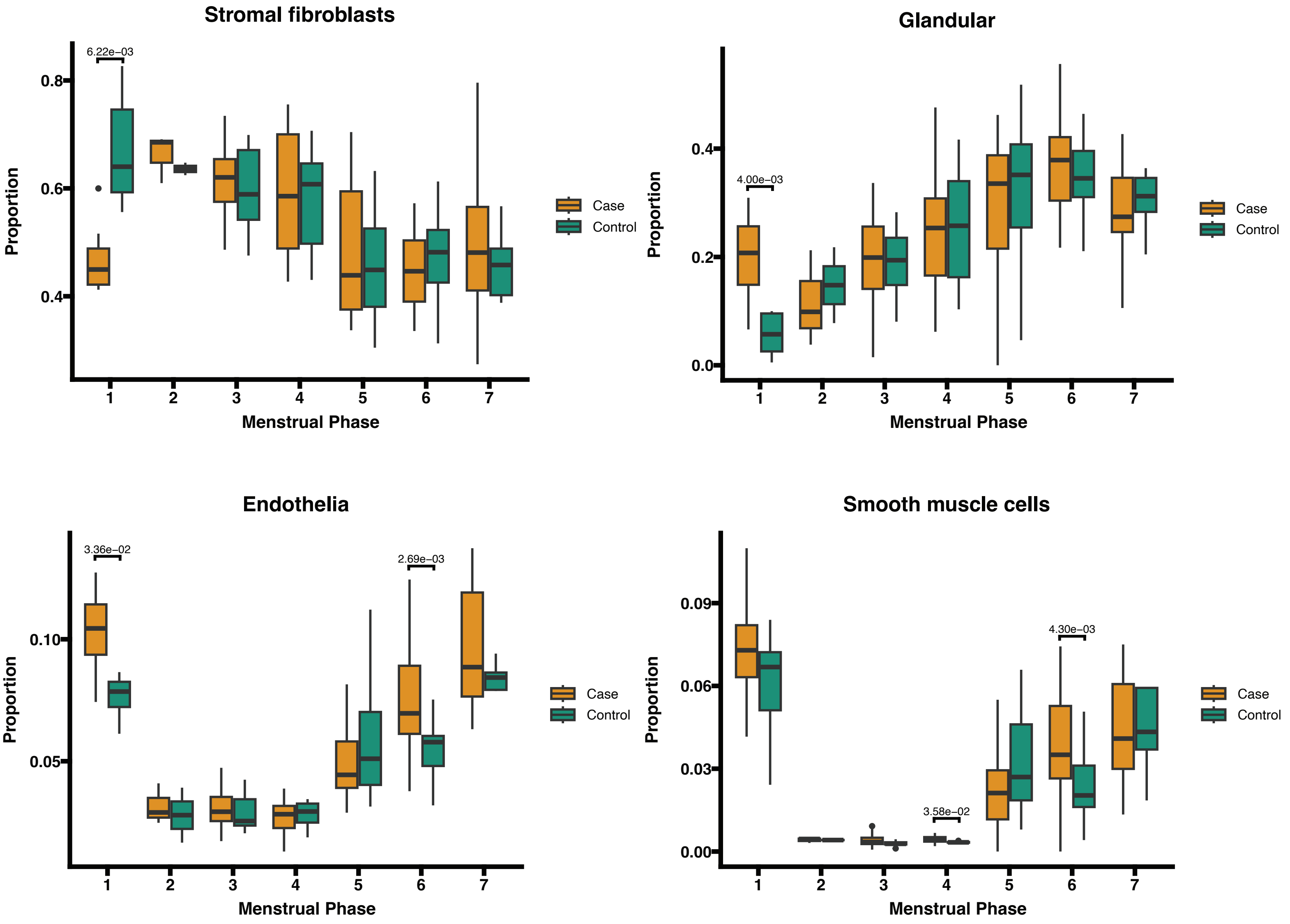


**Figure S2** - Difference between the proportions of cell types between cases (green) and controls (orange) in each of the seven menstrual cycle phases. The phases of the menstrual cycle numbered 1-7 are defined in Figure 2a.

a

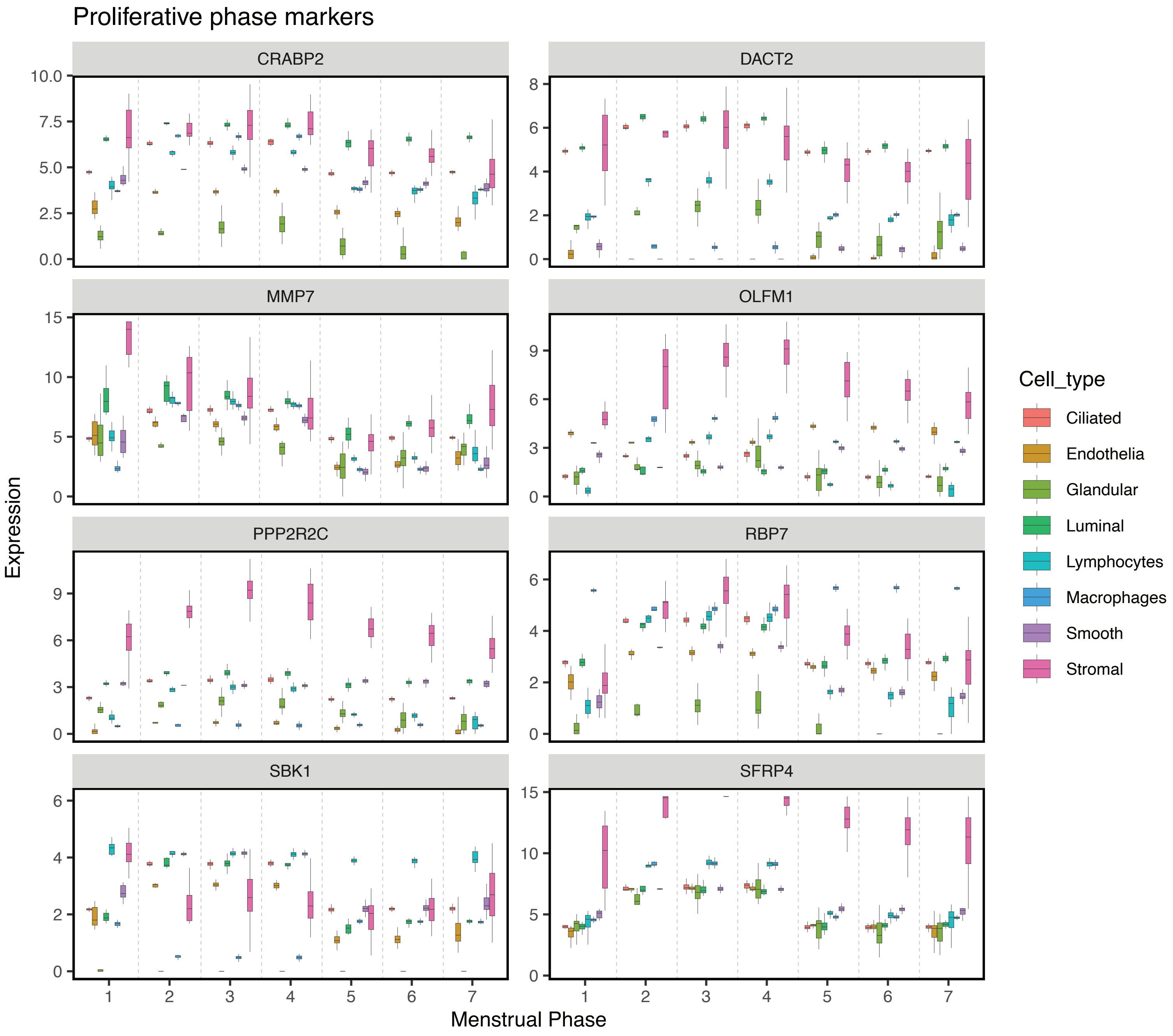

Proliferative phase markers
CRABP2
DACT2
MMP7
OLFM1
PPP2R2C
RBP7
SBK1
SFRP4
Expression
Menstrual Phase
Cell_type
Ciliated
Endothelia
Glandular
Luminal
Lymphocytes
Macrophages
Smooth
Stromal

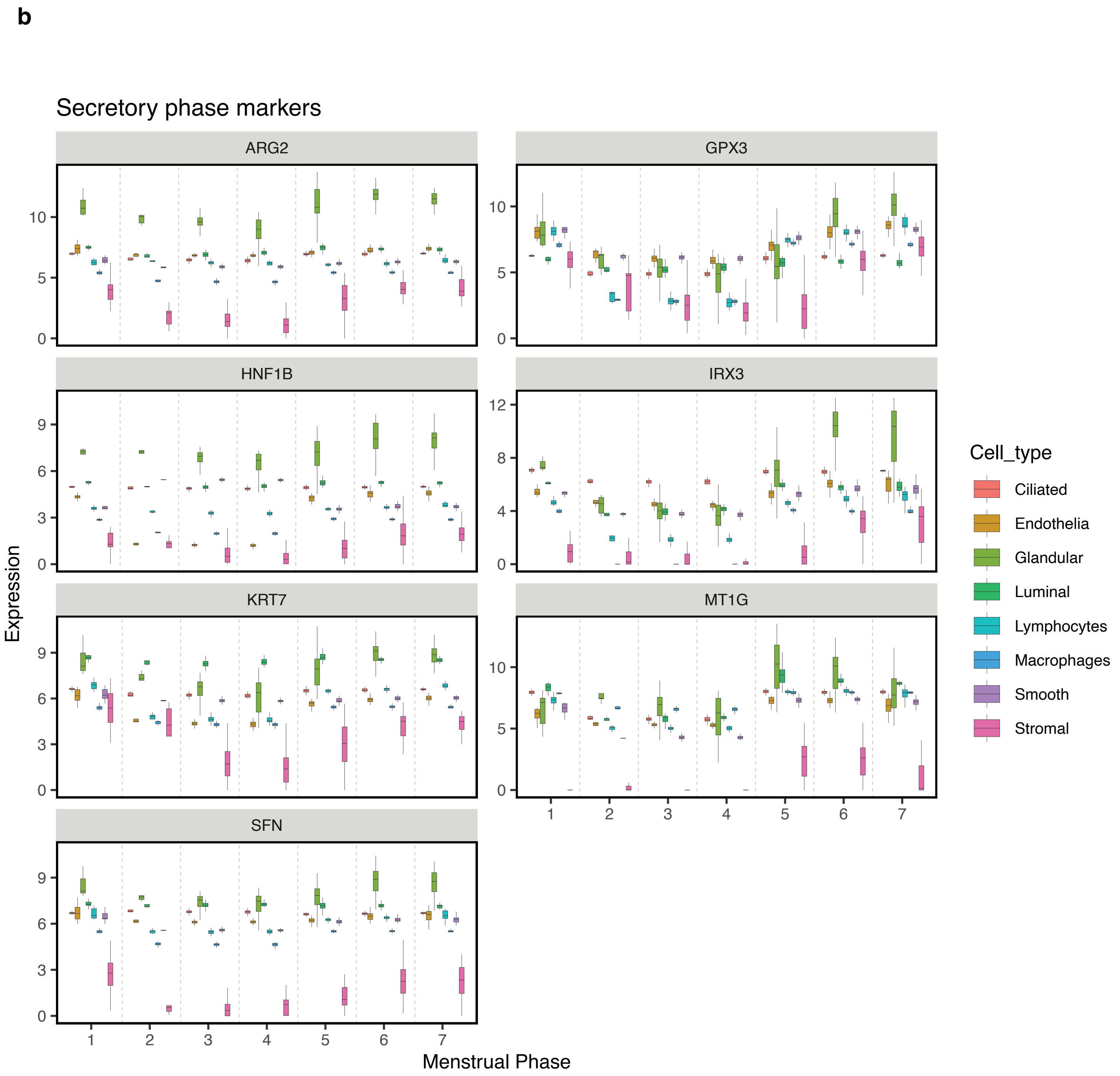


**Figure S3 -** Deconvolution tool bMIND estimated gene expression levels of menstrual phase specific genes which are expressed in higher levels in one menstrual phase compared to the other phase in all cell types. (a) Proliferative phase markers, (b) Secretory phase markers.

**a** Proliferative phase

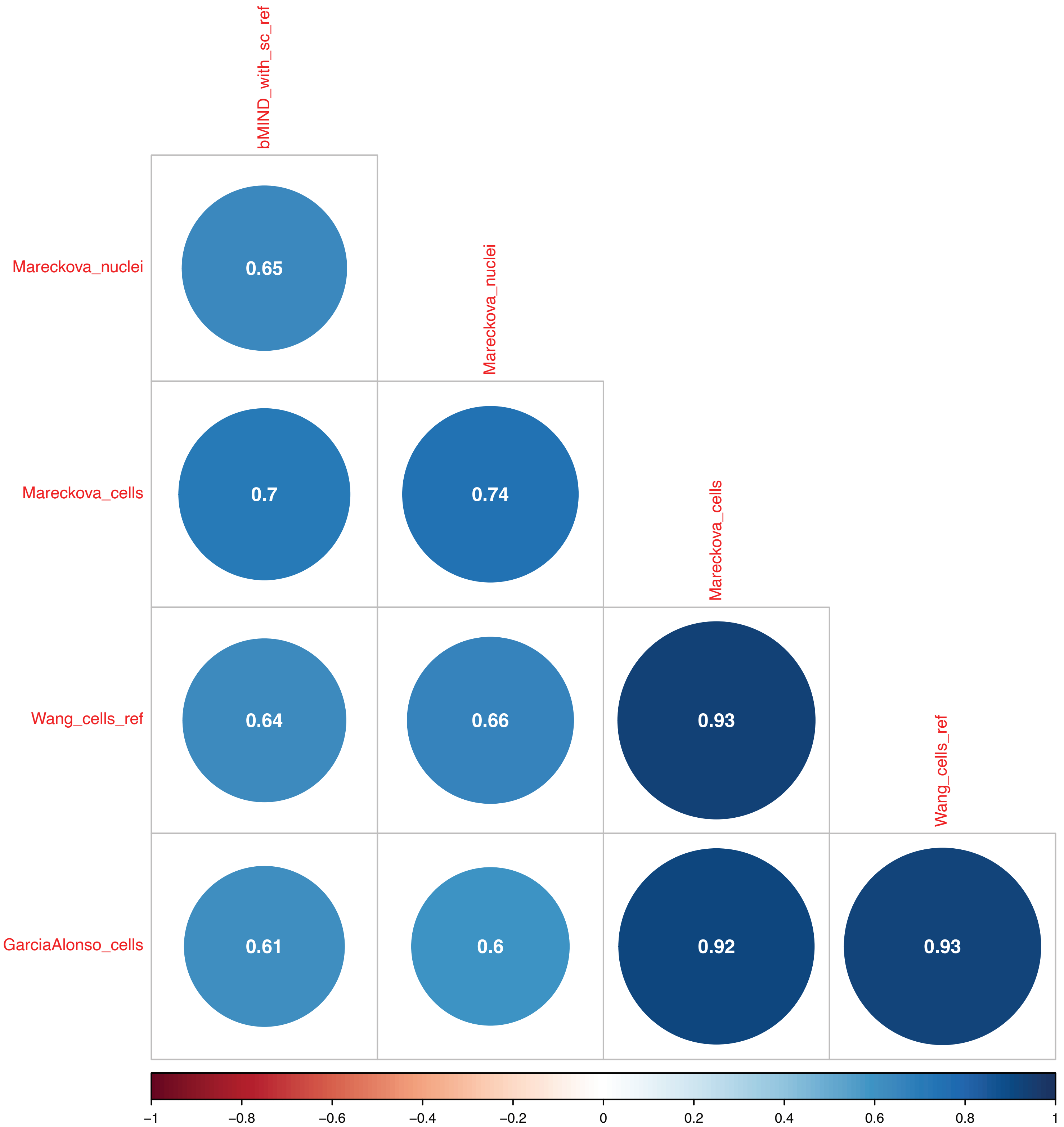

**b** Secretory phase

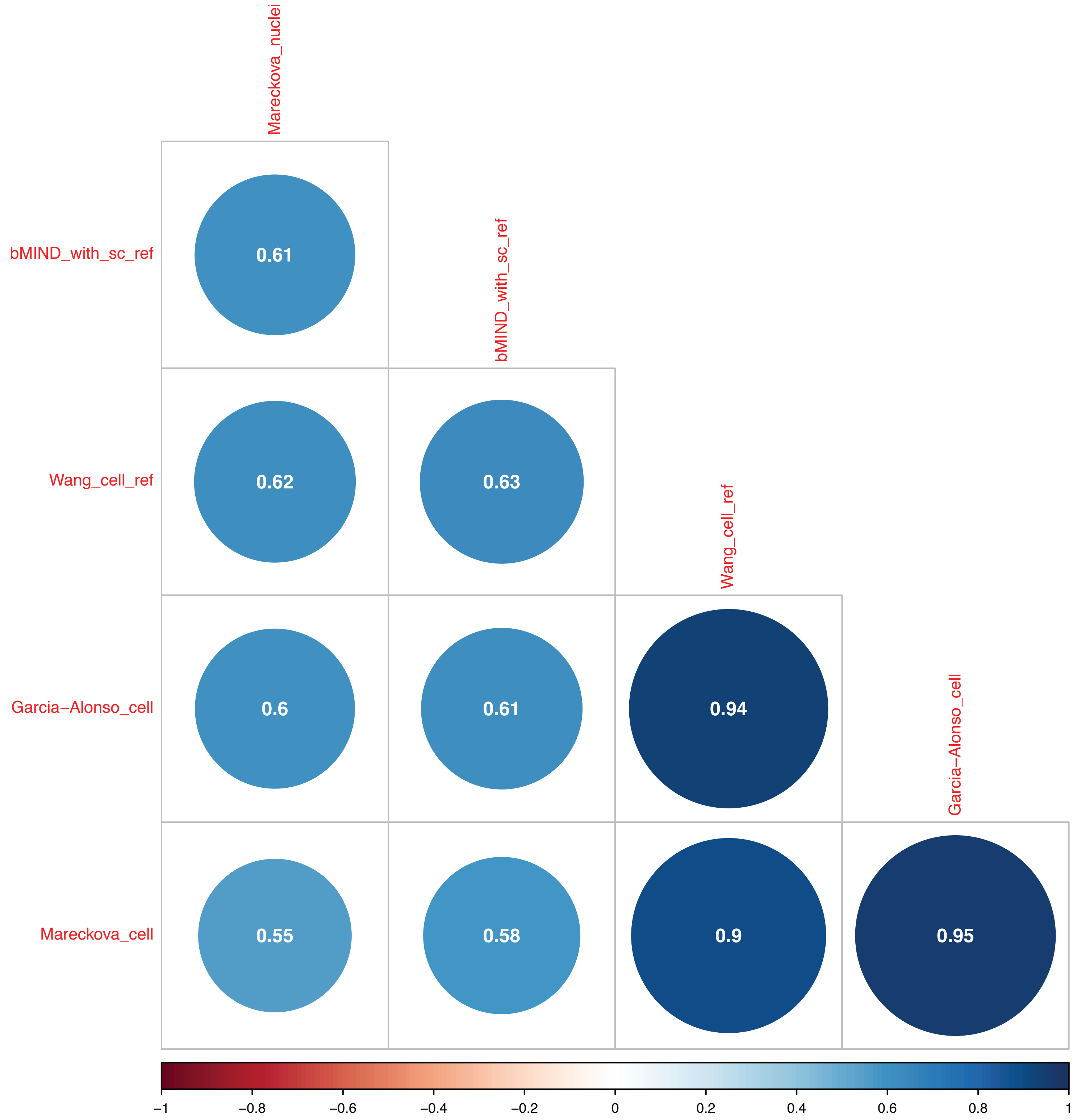


**Figure S4 -** (a) Proliferative phase samples gene expression correlation for stromal fibroblast cell type. (b) Secretory phase samples gene expression correlation for stromal fibroblast cell type. Gene expression correlation between deconvoluted cell type specific estimates and publicly available sc and sn data sets. Label “bMIND_with_sc_ref” refers to the deconvoluted dataset obtained with use of sc data as reference. Other labels correspond to sc dataset names. (a) Proliferative phase comparisons, (b) Secretory phase comparisons.

a

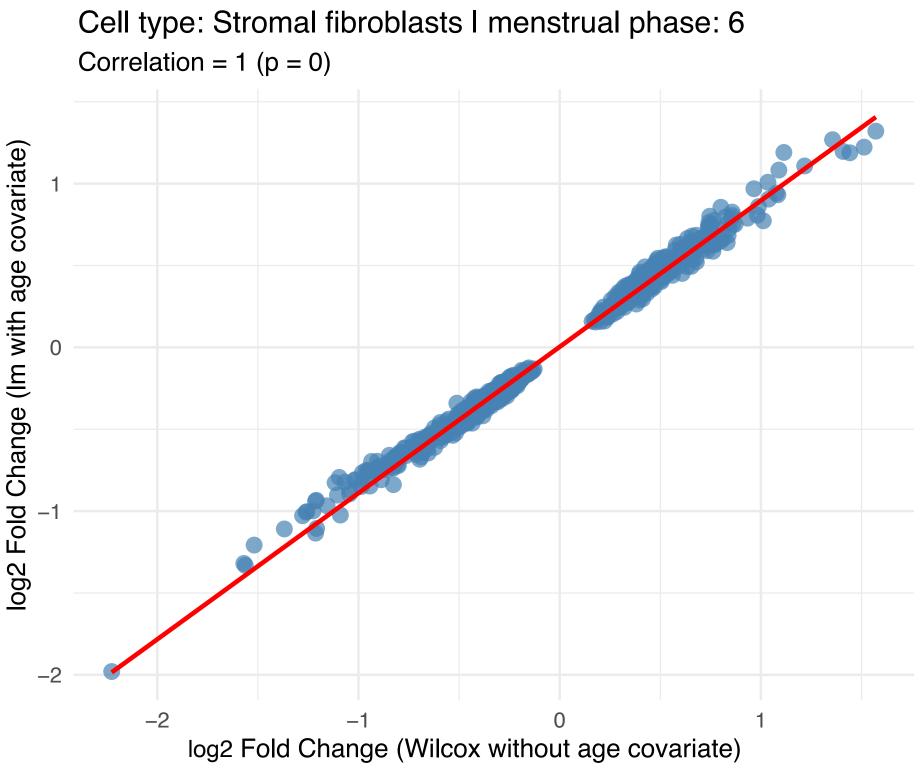


b

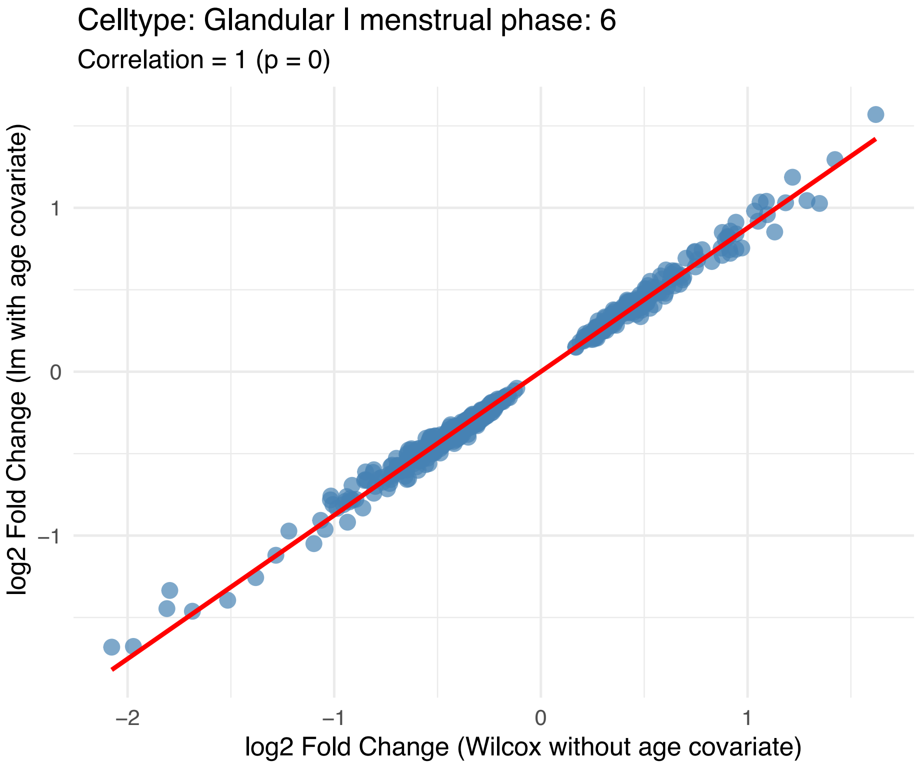


**Figure S5 -** Scatter plots comparing log2 fold changes of differentially expressed genes (FDR < 0.05 in wilcox test results) using two methods: wilcox test without adjusting for age vs linear model with adjusting for age in mid secreotory phase in (a) stromal fibroblasts and (b) glandular epithelial cell types.

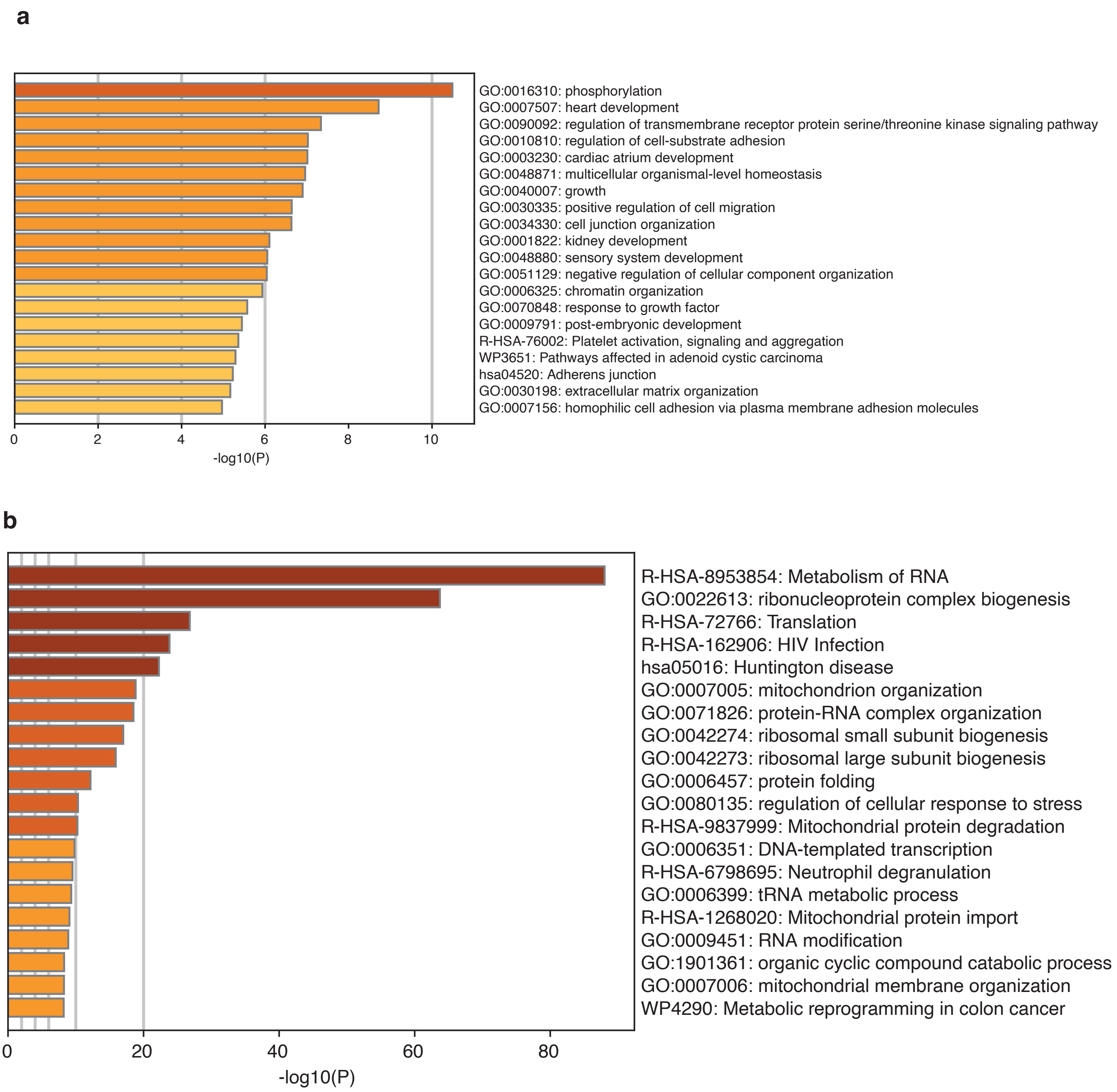
a
GO:0016310: phosphorylation
GO:0007507: heart development
GO:0090092: regulation of transmembrane receptor protein serine/threonine kinase signaling pathway
GO:0010810: regulation of cell-substrate adhesion
GO:0003230: cardiac atrium development
GO:0048871: multicellular organismal-level homeostasis
GO:0040007: growth
GO:0030335: positive regulation of cell migration
GO:0034330: cell junction organization
GO:0001822: kidney development
GO:0048880: sensory system development
GO:0051129: negative regulation of cellular component organization
GO:0006325: chromatin organization
GO:0070848: response to growth factor
GO:0009791: post-embryonic development
R-HSA-76002: Platelet activation, signaling and aggregation
WP3651: Pathways affected in adenoid cystic carcinoma
hsa04520: Adherens junction
GO:0030198: extracellular matrix organization
GO:0007156: homophilic cell adhesion via plasma membrane adhesion molecules
0
2
4
6
8
10
-log10(P)
b
R-HSA-8953854: Metabolism of RNA
GO:0022613: ribonucleoprotein complex biogenesis
R-HSA-72766: Translation
R-HSA-162906: HIV Infection
hsa05016: Huntington disease
GO:0007005: mitochondrion organization
GO:0071826: protein-RNA complex organization
GO:0042274: ribosomal small subunit biogenesis
GO:0042273: ribosomal large subunit biogenesis
GO:0006457: protein folding
GO:0080135: regulation of cellular response to stress
R-HSA-9837999: Mitochondrial protein degradation
GO:0006351: DNA-templated transcription
R-HSA-6798695: Neutrophil degranulation
GO:0006399: tRNA metabolic process
R-HSA-1268020: Mitochondrial protein import
GO:0009451: RNA modification
GO:1901361: organic cyclic compound catabolic process
GO:0007006: mitochondrial membrane organization
WP4290: Metabolic reprogramming in colon cancer
0
20
40
60
80
-log10(P)

**c**

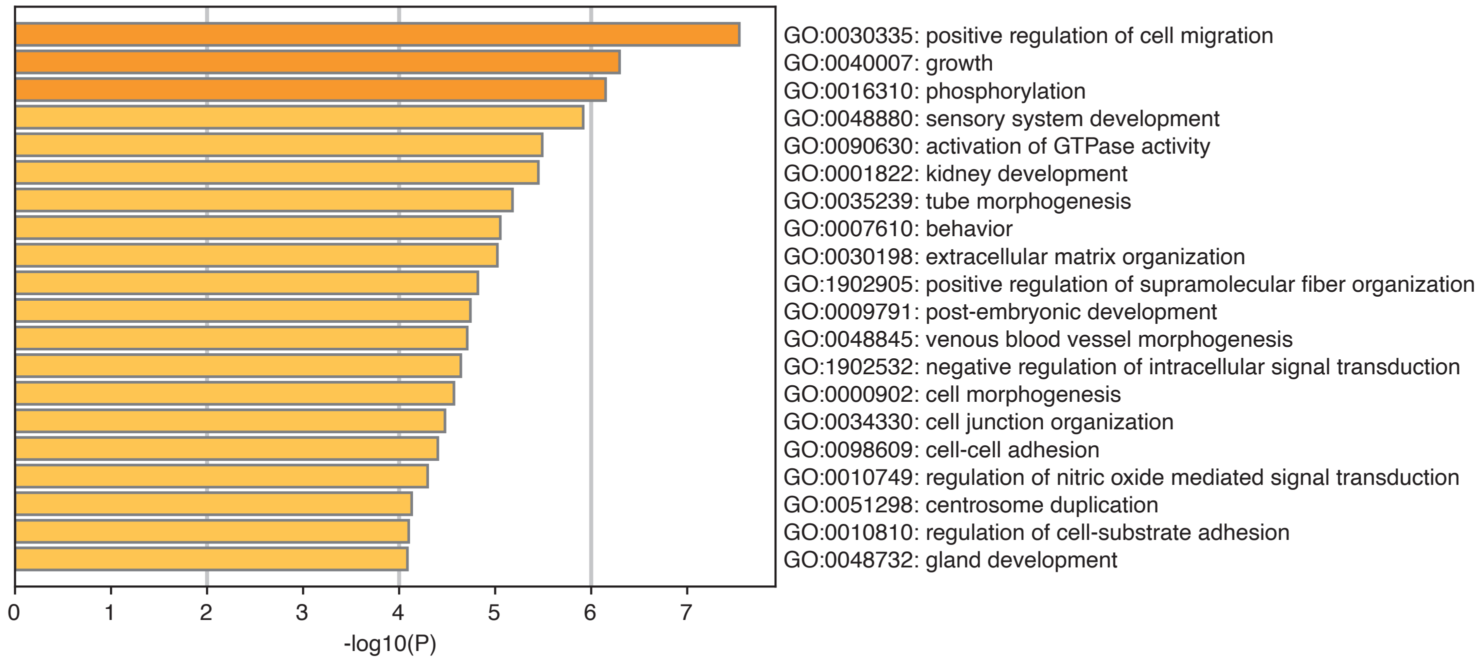
GO:0030335: positive regulation of cell migration
GO:0040007: growth
GO:0016310: phosphorylation
GO:0048880: sensory system development
GO:0090630: activation of GTPase activity
GO:0001822: kidney development
GO:0035239: tube morphogenesis
GO:0007610: behavior
GO:0030198: extracellular matrix organization
GO:1902905: positive regulation of supramolecular fiber organization
GO:0009791: post-embryonic development
GO:0048845: venous blood vessel morphogenesis
GO:1902532: negative regulation of intracellular signal transduction
GO:0000902: cell morphogenesis
GO:0034330: cell junction organization
GO:0098609: cell-cell adhesion
GO:0010749: regulation of nitric oxide mediated signal transduction
GO:0051298: centrosome duplication
GO:0010810: regulation of cell-substrate adhesion
GO:0048732: gland development
0
1
2
3
4
5
6
7
-log10(P)

**d**

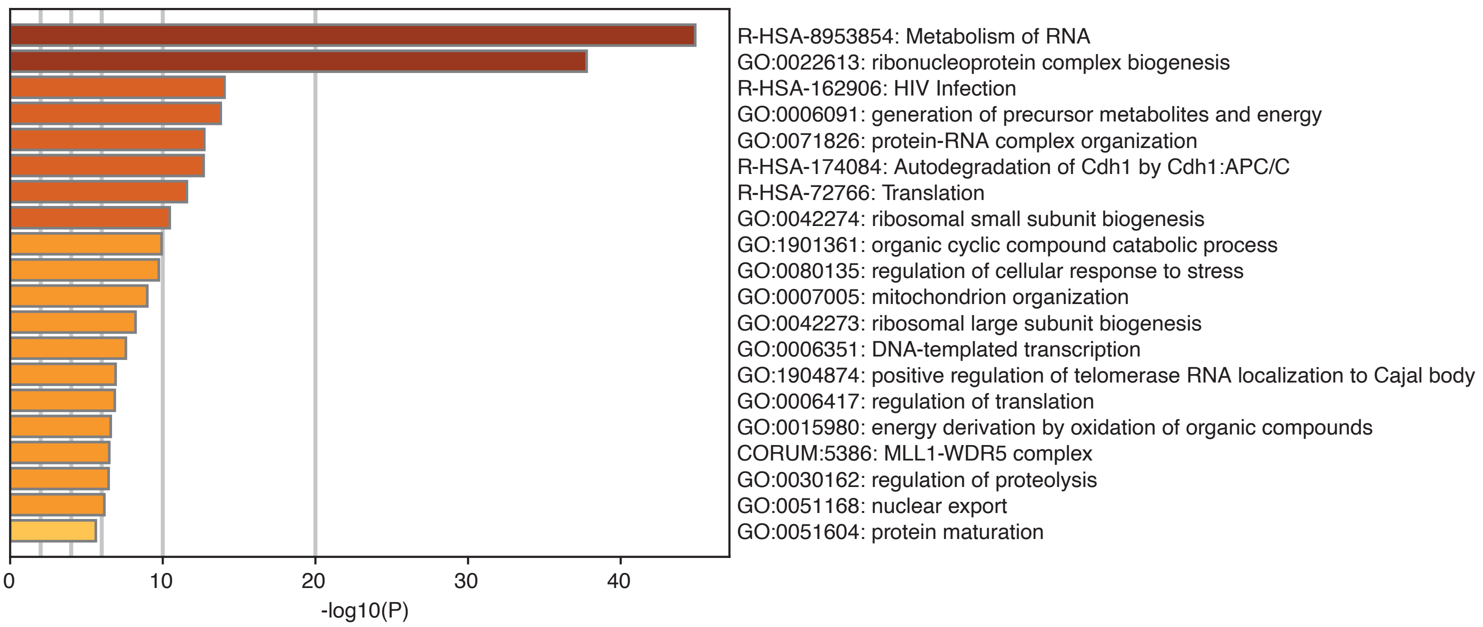
R-HSA-8953854: Metabolism of RNA
GO:0022613: ribonucleoprotein complex biogenesis
R-HSA-162906: HIV Infection
GO:0006091: generation of precursor metabolites and energy
GO:0071826: protein-RNA complex organization
R-HSA-174084: Autodegradation of Cdh1 by Cdh1:APC/C
R-HSA-72766: Translation
GO:0042274: ribosomal small subunit biogenesis
GO:1901361: organic cyclic compound catabolic process
GO:0080135: regulation of cellular response to stress
GO:0007005: mitochondrion organization
GO:0042273: ribosomal large subunit biogenesis
GO:0006351: DNA-templated transcription
GO:1904874: positive regulation of telomerase RNA localization to Cajal body
GO:0006417: regulation of translation
GO:0015980: energy derivation by oxidation of organic compounds
CORUM:5386: MLL1-WDR5 complex
GO:0030162: regulation of proteolysis
GO:0051168: nuclear export
GO:0051604: protein maturation
0
10
20
30
40
-log10(P)

e

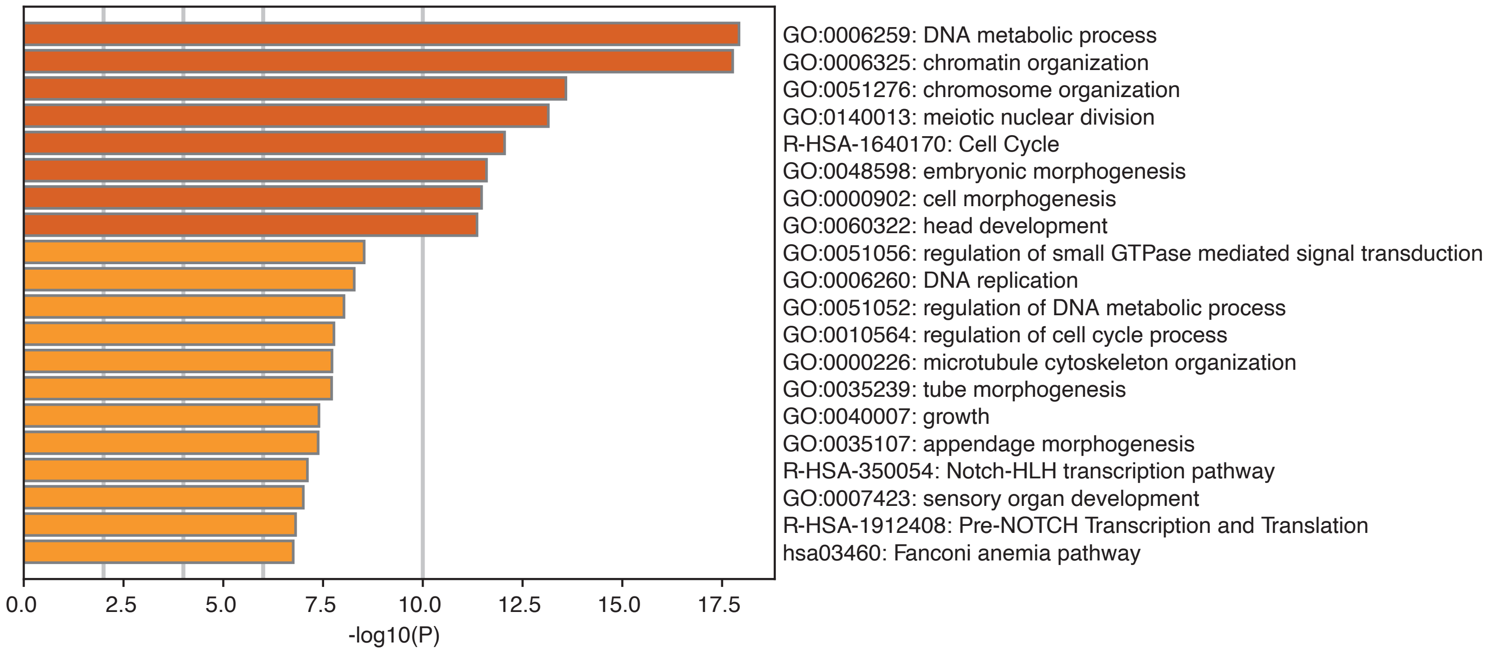


f

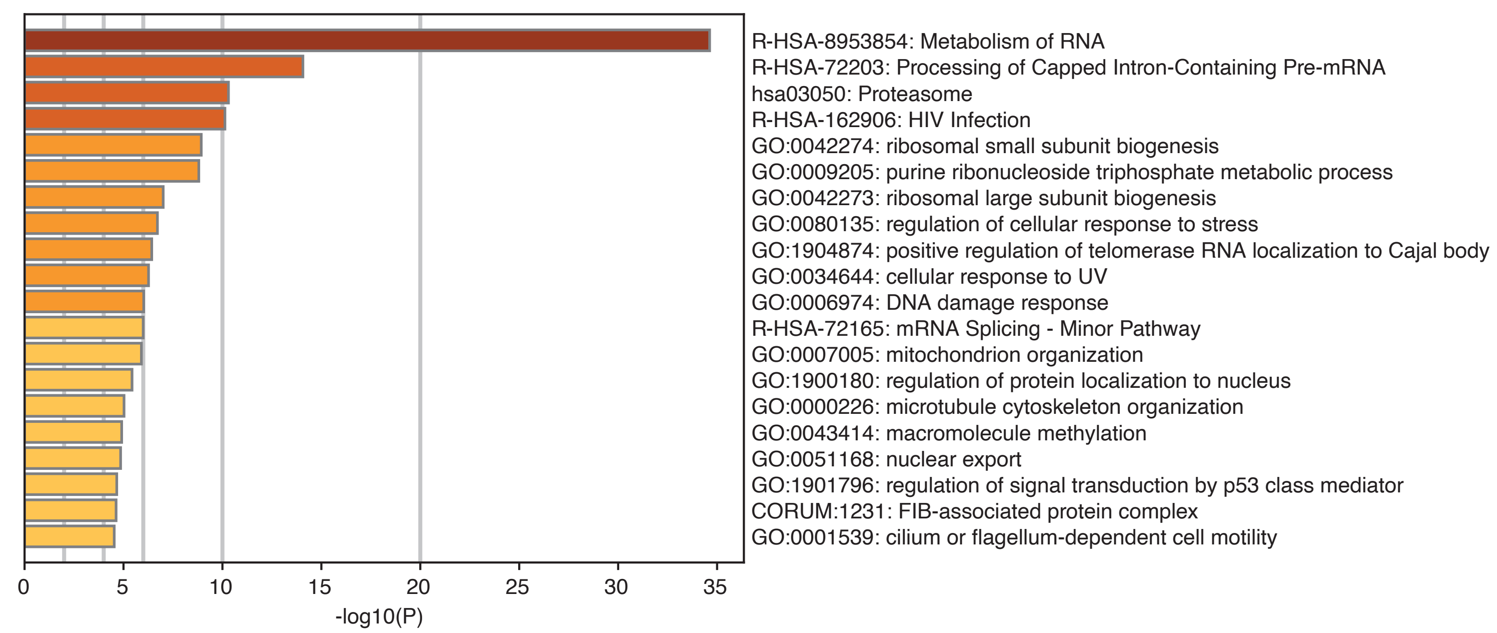


**Figure S6** - Pathways enriched in up and down regulated genes in endometriosis cases in different cell types at the mid secretory phase. Top 20 pathways from each gene set enrichment analysis are shown in figures. (a) Stromal fibroblasts - Up-reulgated DE genes enriched pathways, (b) Stromal fibroblasts - Down-reulgated DE genes enriched pathways, (c) Glandular epithelia - Up-regulated DE genes enriched pathways, (d) Glandular epithelia - Down-regulated DE genes enriched pathways, (e) Luminal epithelia - Up-regulated DE genes enriched pathways, (f) Endothelia - Down-regulated DE genes enriched pathways.
